# Estimating Risk of Mechanical Ventilation and Mortality Among Adult COVID-19 patients Admitted to Mass General Brigham: The VICE and DICE Scores

**DOI:** 10.1101/2020.09.14.20194670

**Authors:** Christopher J. Nicholson, Luke Wooster, Haakon H. Sigurslid, Rebecca F. Li, Wanlin Jiang, Wenjie Tian, Christian L. Lino Cardenas, Rajeev Malhotra

## Abstract

**Background:** Risk stratification of COVID-19 patients upon hospital admission is key for their successful treatment and efficient utilization of hospital resources.

**Objective:** To evaluate the risk factors associated with ventilation need and mortality.

**Design, setting and participants:** We established a retrospective cohort of COVID-19 patients from Mass General Brigham hospitals. Demographic, clinical, and admission laboratory data were obtained from electronic medical records of patients admitted to hospital with laboratory-confirmed COVID-19 before May 19^th^, 2020. Using patients admitted to Massachusetts General Hospital (MGH, derivation cohort), multivariable logistic regression analyses were used to construct the Ventilation in COVID Estimator (VICE) and Death in COVID Estimator (DICE) risk scores.

**Measurements:** The primary outcomes were ventilation status and death.

**Results:** The entire cohort included 1042 patients (median age, 64 years; 56.8% male). The derivation and validation cohorts for the risk scores included 578 and 464 patients, respectively. We found seven factors to be independently predictive for ventilation requirement (diabetes mellitus, dyspnea, alanine aminotransferase, troponin, C-reactive protein, neutrophil-lymphocyte ratio, and lactate dehydrogenase), and 10 factors to be predictors of in-hospital mortality (age, sex, diabetes mellitus, chronic statin use, albumin, C-reactive protein, neutrophil-lymphocyte ratio, mean corpuscular volume, platelet count, and procalcitonin). Using these factors, we constructed the VICE and DICE risk scores, which performed with C-statistics of at least 0.8 in our cohorts. Importantly, the chronic use of a statin was associated with protection against death due to COVID-19. The VICE and DICE score calculators have been placed on an interactive website freely available to the public (https://covid-calculator.com/).

**Limitations:** One potential limitation is the modest sample sizes in both our derivation and validation cohorts.

**Conclusion:** The risk scores developed in this study may help clinicians more appropriately determine which COVID-19 patients will need to be managed with greater intensity.

## Introduction

The number of global confirmed cases of the novel coronavirus disease 2019 (COVID-19) passed 20 million in August, with over 700,000 deaths.^1^ The U.S. has surpassed any other country in the number of total deaths and case rates continue to rise with some hospitals utilizing nearly 100% of available ICU beds.

Specific information regarding the patient risk factors that associate with mortality from COVID-19 remain limited, and methods to accurately predict severity of disease at the time of hospital presentation are lacking.^2-6^ Using data from a Chinese cohort, an algorithm was recently developed that predicts critical illness (a composite of ICU admission, ventilation needs and death) in hospitalized COVID-19 patients. However, the applicability of this algorithm to predict outcomes in a US population, which has distinct disease risk profiles, remains unknown.^7,8^ Further, it is crucial for health care providers to be able to stratify risk for the most important clinical outcomes in COVID, namely mechanical ventilation need and mortality. Knowing the risk of these outcomes will allow the most optimal allocation of healthcare resources on admission to the hospital and identify those that will require the most intense care. Given the United States has reported over a quarter of global deaths due to COVID-19 and is currently in the midst of a profound wave of infections, new information on the factors that influence risk of severe outcomes is greatly needed.

This study describes the details of patients with laboratory confirmed COVID-19 admitted to Mass General Brigham hospitals in Boston, Massachusetts. Specifically, we described the baseline comorbidities, presenting clinical tests and outcomes of hospitalized COVID-19 patients, explored the risk factors associated with mechanical ventilation requirements and in-hospital death, and developed risk models to more effectively predict severe outcomes in patients from the United States.

## Methods

### Study Population

This study included consecutive adult patients with laboratory-confirmed COVID-19 infection who were admitted for illness related to COVID-19 to five hospitals in the Mass General Brigham health care system (Massachusetts General Hospital, MGH; Brigham and Women’s Hospital, BWH; Newton Wellesley Hospital, NWH; Brigham and Women’s Faulkner Hospital, BWFH; and North Shore Medical Center, NSMC) in the Boston region before May 19, 2020. Decisions to admit to hospital were made subjectively by the assessing clinicians at point of referral. The Mass General Brigham institutional review board (IRB) approved the study (Protocol # 2020P000982).

A confirmed case of COVID-19 was defined by a positive result on a reverse-transcriptase-polymerase-chain-reaction (RT-PCR) assay of a specimen collected on a nasopharyngeal swab. Patients were defined as COVID-19 positive if they had a positive test, or if they had a negative test, but repeat testing was positive. Only laboratory-confirmed cases of those that were sufficiently ill to require hospital admission were included. Clinical outcomes were monitored up to and including July 20, 2020, the final date for follow-up. We excluded children (those younger than 18 years of age) from the study.

### Data collection

Epidemiological, demographic (self-reported), clinical laboratory, and treatment data were obtained first from the Research Patient Data Registry, a centralized clinical data registry directed by the Mass General Brigham network. Outcome data, including discharge, ICU, and ventilation status, and home medication data were extracted from electronic medical records (EPIC) using a standardized data collection form. All laboratory tests and radiologic assessments, including plain chest radiography, were performed at the discretion of the treating physician. Only laboratory tests performed on or within 24 hours of hospital admission were included in the analyses. Patients were assessed for the presence of hypertension, diabetes mellitus, coronary artery disease, chronic obstructive pulmonary disease (COPD), chronic kidney disease (CKD), and a history of cancer. These covariates were selected based relevance to COVID-19 in previously published analyses. ^2,3,5-7,9^

### Outcome definitions and data analysis

The primary endpoints for our analyses were need for mechanical ventilation and in-hospital death. For our statistical analyses, we excluded patients that requested, upon admission, to be treated with comfort measures only (CMO). Patients that were identified as “do not intubate” (DNI) on admission were excluded from analyses that assessed risk factors for mechanical ventilation requirements. Discharged patients were defined as those who were discharged to home, nursing homes, or a rehabilitation facility. All analyses were performed using R studio (version 1.2.5033) or Stata (version 13.0). Continuous variables were reported as mean (SD) unless otherwise noted and categorical variables were reported as n (%). The Fisher’s exact test, Student’s t-test or Mann Whitney test were used to measure differences between groups, where appropriate. Univariate logistic regression was used to determine if a clinical factor was associated with the need for mechanical ventilation or with in-hospital mortality. Validation was performed by applying the regression coefficients and estimating probabilities.

### Multivariable logistic regression and risk score construction

We used a multivariable logistic regression model to determine variables that would be included in our predictive risk score algorithms for mechanical ventilation needs (Ventilation in COVID Estimator [VICE] score) and death (Death in COVID Estimator [DICE] score). To do so, we used a derivation cohort including patients admitted to MGH only. For variables that had a univariate P-value of <0.05 or a P-value <0.05 after adjusting for age and sex in this cohort, we performed a multivariable logistic regression analysis with a backwards stepwise approach. Of variables that were highly correlative (e.g. eGFR and creatinine, with r > 0.7), we only included the one with the lowest p-value in univariate analysis. We used this method to determine risk factors associated with (i) mechanical ventilation requirements and (ii) in-hospital mortality, which allowed construction of risk score predictors for each outcome. We assessed the accuracy of the risk score models using the area under the receiver-operator characteristic curve (AUC or C-statistic). We first assessed our risk score in patients that were admitted to MGH only (our derivation cohort), and then validated it using patients admitted to BWH, NWH, BWFH, and NSMC (our validation cohort).

## Results

### Demographic and clinical characteristics of the cohort

1137 adults were admitted to Mass General Brigham hospitals with COVID-19 symptoms before May 19, 2020. Patients that were treated with comfort measures only (CMO) on arrival (n=95) to the hospital were excluded from the study. As described in Supplemental Table 3, CMO patients were on average older and had a higher level of cancer diagnoses than patients included in the final study. Interestingly, however, only 17% of these patients died in hospital by the end of the study. After this exclusion, we included 1042 patients (578 from MGH, 269 from BWH, 125 from BWFH, 60 from NWH, and 10 from NSMC) in our final analyses (**Table 1**). The median age for these patients was 64 (IQR 53-75), ranging from 18 to 99 years old, and the majority (57%) were male. Among the 1042 patients, 438 (42%) identified as white, 187 (17.9%) as black, 113 (10.8%) as Hispanic, and 37 (3.6%) as Asian. One hundred and seventy-seven (17%) patients did not identify with any of these racial backgrounds and 90 (8.6%) patients had no racial background recorded.

**Table 1:** Baseline characteristics of hospitalized Covid-19 patients included in the study.

| Characteristic | All patients | Ventilation Status |  |  |  | Mortality |  |  |  |
| --- | --- | --- | --- | --- | --- | --- | --- | --- | --- |
|  |  | Not ventilated | Ventilated | OR (95% CI) | P-Value | Discharged | Deceased | OR (95% CI) | P-Value |
| Total patients | 1042 | 550 | 401 |  | - | 829 | 211 |  | - |
| Median (IQ) Age | 64 (53-75) | 61 (51-72) | 64 (53-72) | 1.07 (0.99-1.16)<sup>∞</sup> | 0.104 | 61 (50-71) | 75 (66-82) | 1.99 (1.75-2.27)<sup>∞</sup> | <0.001 |
| Male Sex (%) | 592 (56.8) | 285 (51.8) | 263 (65.6) | 1.77 (1.36-2.31) | <0.001 | 446 (54.1) | 140 (66.4) | 1.66 (1.21-2.28) | 0.002 |
| **Race (%\*)** |  |  |  |  |  |  |  |  |  |
| White | 438 (42.0) | 220 (59.1) | 152 (40.9) | - | - | 321 (73.3) | 117 (26.7) | - | - |
| Black | 187 (17.9) | 110 (62.9) | 65 (37.1) | 0.92 (0.77-1.11) | 0.407 | 149 (80.1) | 37 (19.9) | 0.83 (0.67-1.02) | 0.072 |
| Hispanic | 113 (10.8) | 71 (65.1) | 38 (34.9) | 0.77 (0.50-1.21) | 0.261 | 96 (85.0) | 17 (15.0) | 0.49 (0.28-0.85) | 0.011 |
| Asian | 37 (3.6) | 15 (45.5) | 18 (54.5) | 1.20 (0.95-1.53) | 0.131 | 32 (88.9) | 4 (11.1) | 0.70 (0.49-0.997) | 0.048 |
| Other/Mix | 177 (17.0) | 98 (56.0) | 77 (44.0) | 1.03 (0.94-1.13) | 0.546 | 159 (89.8) | 18 (10.2) | 0.75 (0.66-0.86) | <0.001 |
| Not recorded | 90 (8.6) | 36 (41.4) | 51 (58.6) | - | - | 72 (8.7) | 18 (8.5) | - | - |
| **Comorbidity (%)** |  |  |  |  |  |  |  |  |  |
| Diabetes | 443 (42.5) | 203 (36.9) | 194 (48.4) | 1.60 (1.23-2.07) | <0.001 | 330 (39.8) | 111 (52.6) | 1.70 (1.25-2.30) | 0.001 |
| Coronary artery disease | 182 (17.5) | 85 (15.5) | 60 (15.0) | 0.96 (0.67-1.38) | 0.826 | 123 (14.8) | 59 (28) | 2.24 (1.57-3.20) | <0.001 |
| Hypertension | 588 (56.4) | 295 (53.6) | 224 (55.9) | 1.09 (0.84-1.41) | 0.516 | 444 (53.6) | 142 (67.3) | 1.81 (1.32-2.49) | <0.001 |
| Hyperlipidemia | 397 (38.1) | 211 (38.4) | 141 (35.2) | 0.87 (0.66-1.14) | 0.303 | 299 (36.1) | 98 (46.4) | 1.55 (1.14-2.11) | 0.005 |
| Chronic kidney disease | 174 (16.7) | 87 (15.8) | 56 (14.0) | 0.86 (0.60-1.24) | 0.423 | 114 (13.8) | 59 (28.0) | 2.45 (1.71-3.51) | <0.001 |
| COPD | 123 (11.8) | 54 (9.8) | 44 (11.0) | 1.13 (0.74-1.72) | 0.57 | 77 (9.3) | 45 (21.3) | 2.66 (1.78-3.99) | <0.001 |
| Cancer | 166 (15.9) | 89 (16.2) | 47 (11.7) | 0.68 (0.46-0.99) | 0.045 | 113 (13.6) | 53 (25.1) | 2.12 (1.47-3.07) | <0.001 |
| **Symptom (%)** |  |  |  |  |  |  |  |  |  |
| Dyspnea | 739 (70.9) | 343 (62.4) | 339 (84.5) | 3.31 (2.39-4.58) | <0.001 | 587 (70.8) | 150 (71.1) | 1.04 (0.74-1.46) | 0.821 |
| LOC | 30 (2.9) | 20 (3.6) | 6 (1.5) | 0.40 (0.16-1.00) | 0.05 | 22 (2.7) | 8 (3.8) | 1.46 (0.64-3.32) | 0.371 |
| Hemoptysis | 13 (1.2) | 7 (1.3) | 5 (1.2) | 0.97 (0.31-3.07) | 0.957 | 12 (1.5) | 1 (0.5) | 0.33 (0.04-2.52) | 0.282 |
| **Smoking (%\*)** |  |  |  |  |  |  |  |  |  |
| Never | 387 (37.1) | 211 (57.7) | 155 (42.3) | - | - | 320 (83.1) | 65 (16.9) | - | - |
| Former | 231 (22.2) | 106 (53.5) | 92 (46.5) | 1.18 (0.83-1.67) | 0.347 | 163 (70.6) | 68 (29.4) | 2.05 (1.39-3.03) | <0.001 |
| Current | 86 (8.3) | 37 (47.4) | 41 (52.6) | 1.23 (0.96-1.57) | 0.100 | 70 (81.4) | 16 (18.6) | 1.06 (0.78-1.44) | 0.702 |
| Not Recorded | 338 (32.4) | 196 (63.4) | 113 (36.6) | - | - | 276 (81.7) | 62 (18.3) | - | - |
| **Medication (%)** |  |  |  |  |  |  |  |  |  |
| Statin | 511 (49.0) | 270 (49.1) | 179 (44.6) | 0.84 (0.65-1.09) | 0.201 | 388 (46.8) | 122 (57.8) | 1.61 (1.18-2.18) | 0.003 |
| RAAS inhibitor | 315 (30.2) | 163 (29.6) | 125 (31.2) | 1.08 (0.81-1.42) | 0.611 | 247 (29.8) | 67 (31.8) | 1.10 (0.79-1.52) | 0.58 |
| Aspirin | 318 (30.5) | 170 (30.9) | 106 (26.4) | 0.81 (0.61-1.08) | 0.155 | 232 (28.0) | 86 (40.8) | 1.80 (1.32-2.47) | <0.001 |
| **Abnormal X-ray** | 825 (79.2) | 389 (70.7) | 363 (90.5) | 3.81 (2.55-5.70) | <0.001 | 641 (77.3) | 175 (82.9) | 1.44 (0.94-2.18) | 0.091 |

Patients coded for “do not intubate” were excluded from analyses for ventilation status. P values are for univariate logistic regression analyses. COPD = chronic obstructive pulmonary disorder; LOC = loss of consciousness; RAAS = renin angiotensin-aldosterone system; OR = odds ratio; CI = confidence interval; SD = standard deviation.
\* Calculated as a percentage of each group (race or smoking status). Otherwise, values were calculated as a percentage of each outcome group (ventilation or discharge). Odds ratios in the ethnicity and smoking categories were calculated relative to white patients or never smokers, respectively. <sup>∞</sup> Odds ratio calculated for every 10-year increase in age.

As of July 20th, 2020, among the 1042 patients admitted to hospital, 829 (79.6%) were discharged, 211 (20.2%) died in hospital, and 2 (0.2%) remained in hospital. Three quarters of the patients had at least one comorbidity. The most common comorbidities were hypertension (56.4%), diabetes mellitus (42.5%), and hyperlipidemia (38.1%). 86% of patients who died had at least one comorbidity. The median length of stay was 10 (IQR: 6-21, up to 98) days. The median length of ICU stay (n=449) and ventilation time (n=400) were 15 (7-23) days and 13 (7-22) days, respectively. The median survival time amongst those that died was 11 days (IQR = 6-19; Range = 1-71). Only 47 patients who were admitted to the ICU received no mechanical ventilation. Ninety-one patients were identified as “do not intubate” (DNI) on admission. As shown in Supplemental Table 3, DNI patients were on average older and had a higher prevalence of pre-existing conditions than patients included in the final study. Of the 449 patients who were admitted to the ICU, 400 patients (89.1%) required mechanical ventilation. One hundred and thirty-six (34%) mechanically ventilated patients died. With regards to home medications, 511 (49%) were on a statin, 318 (30.5%) on aspirin, and 315 (30.2%) on a RAAS inhibitor.

### Patient factors associated with severity of disease

In separate univariate analyses, clinical factors that were associated with need for mechanical ventilation and associated with in-hospital mortality were identified (detailed in **Table 1** and **Table 2**). Many variables on admission were consistently predictive of both ventilation need and mortality, including male sex, diabetes mellitus, lower levels of albumin and eGFR, and elevated absolute neutrophils, anion gap, activated partial thromboplastin time, blood urea nitrogen, C-reactive protein, creatinine, D-dimer, eGFR, plasma glucose, neutrophil to lymphocyte ratio, procalcitonin, and troponin T (high sensitivity). Troponin predicted both need for mechanical ventilation and in-hospital mortality when assessed as a continuous variable or as a dichotomous variable with the threshold of >10 ng/mL used to indicate the presence of myocardial injury. However, it was striking that many factors were only associated with only one of either mechanical ventilation need or in-hospital mortality. Those that met significance for ventilation need only included dyspnea and loss of consciousness on presentation, x-ray abnormality on admission, and elevated alanine aminotransferase (ALT), aspartate aminotransferase (AST), direct bilirubin, erythrocyte sedimentation rate (ESR), ferritin, fibrinogen, lactate dehydrogenase (LDH), mean corpuscular hemoglobin (MCH), and white blood cell count (WBC). The variables that were predictive of mortality only included age, coronary artery disease (CAD), hypertension, hyperlipidemia, chronic kidney disease, chronic obstructive pulmonary disease, statin use, aspirin use, lower levels of hemoglobin and platelets, and elevated levels of creatinine, mean corpuscular volume (MCV), NT-ProBNP, and increased prothrombin time, and red cell distribution and width (RDW).

**Table 2:** Laboratory results of hospitalized COVID-19 patients on admission.

|  |  | Ventilation Status |  |  | Mortality |  |  |
| --- | --- | --- | --- | --- | --- | --- | --- |
| Lab test | Mean (SD)<br>Total | Mean (SD)<br>Not ventilated<br>(n = 550) | Mean (SD)<br>Ventilated<br>(n = 401) | P-Value | Mean (SD)<br>Discharged<br>(n = 829) | Mean (SD)<br>Deceased<br>(n = 211) | P-Value |
| Absolute Lymphocytes (K/μL) | 1.36 (5.38) | 1.38 (5.37) | 1.35 (5.87) | 0.931 | 1.31 (4.70) | 1.55 (7.51) | 0.574 |
| Absolute Neutrophils (K/μL) | 5.98 (3.98) | **5.28 (3.91)** | **6.92 (3.69)** | **<0.001** | **5.80 (3.90)** | **6.66 (4.24)** | **0.009** |
| Albumin (g/dL) | 3.62 (0.52) | **3.74 (0.47)** | **3.47 (0.53)** | **<0.001** | **3.68 (0.50)** | **3.38 (0.54)** | **<0.001** |
| Anion Gap (mmol/L) | 16.3 (3.6) | **15.9 (3.6)** | **16.8 (3.7)** | **<0.001** | **16.2 (3.4)** | **16.8 (4.2)** | **0.02** |
| Alanine aminotransferase (U/L) | 52.5 (234.9) | **37.2 (40.3)** | **77.3 (371.2)** | **<0.001** | 47.7 (111.2) | 71.0 (473.2) | 0.277 |
| aPTT (s) | 36.8 (14.5) | **34.7 (11.2)** | **38.6 (17.0)** | **0.01** | **35.9 (12.5)** | **40.4 (19.9)** | **0.009** |
| Aspartate aminotransferase (U/L) | 79.5 (492.4) | **48.5 (59.5)** | **128.8 (786.7)** | **<0.001** | 65.5 (217.1) | 134.3 (1006) | 0.195 |
| Blood Urea Nitrogen (mg/dL) | 22.2 (18.7) | **19.9 (16.8)** | **23.8 (20.6)** | **0.002** | **19.4 (16.2)** | **33.3 (23.2)** | **<0.001** |
| C-reactive protein (mg/L) | 109.6 (88.4) | **79.9 (70.0)** | **154.9 (94.8)** | **<0.001** | **102.6 (86.4)** | **136.3 (91.5)** | **<0.001** |
| Creatinine (mg/dL) | 1.48 (1.87) | **1.35 (1.52)** | **1.61 (2.30)** | **0.049** | **1.35 (1.82)** | **1.99 (2.00)** | **<0.001** |
| Creatine Kinase (U/L) | 764.0 (11178) | 978.3 (15267) | 581.2 (2538) | 0.64 | 793.8 (12494) | 642.2 (2252) | 0.868 |
| D-dimer (ng/L) | 1905 (3327) | **1500 (1513)** | **2370 (4880)** | **<0.001** | **1748 (3514)** | **2489 (2321)** | **0.044** |
| Direct bilirubin (mg/dL) | 0.22 (0.42) | **0.17 (0.36)** | **0.28 (0.51)** | **0.001** | 0.21 (0.42) | 0.25 (0.38) | 0.238 |
| eGFR (mL/min/1.73m<sup>2</sup>) | 67.9 (29.8) | **71.4 (29.1)** | **66.8 (30.5)** | **0.02** | **72.6 (28.4)** | **49.3 (27.8)** | **<0.001** |
| ESR (mm/hr) | 50.3 (31.1) | **47.9 (29.8)** | **54.8 (32.6)** | **0.004** | 49.8 (30.4) | 52.8 (33.8) | 0.281 |
| Ferritin (μg/mL) | 1283 (3872) | **812.8 (1116)** | **2037 (5988)** | **<0.001** | 1173 (3540) | 1688 (4971) | 0.137 |
| Fibrinogen (mg/dL) | 578.3 (175.9) | **529.2 (149.1)** | **638.5 (187.3)** | **<0.001** | 583.5 (178.2) | 556.7 (166.7) | 0.214 |
| Glucose (mg/dL) | 157.4 (94.63) | **148.1 (96.24)** | **172.0 (96.24)** | **<0.001** | **152.6 (91.74)** | **176.6 (103.6)** | **0.002** |
| Hemoglobin (g/dl) | 12.9 (2.1) | 12.9 (2.0) | 13.0 (2.2) | 0.212 | **13.0 (2.0)** | **12.3 (2.5)** | **<0.001** |
| Lactate Dehydrogenase (U/L) | 408.4 (558.6) | **323.8 (153.4)** | **530.6 (846.8)** | **<0.001** | 384.2 (310.0) | 504.0 (1083) | 0.102 |
| MCH (pg/cell) | 29.0 (2.5) | **28.8 (2.6)** | **29.2 (2.3)** | **0.016** | 28.9 (2.5) | 29.3 (2.4) | 0.056 |
| MCV (fL/cell) | 87.9 (6.5) | 87.4 (6.7) | 88.1 (6.1) | 0.091 | **87.4 (6.3)** | **89.8 (7.00)** | **<0.001** |
| Neut:Lymph Ratio | 8.92 (13.23) | **6.73 (8.86)** | **11.54 (16.40)** | **<0.001** | **7.66 (9.88)** | **13.83 (21.28)** | **<0.001** |
| NT_Pro-BNP (pg/mL) | 3233 (12280) | 2648 (13728) | 3507 (10668) | 0.449 | **2339 (12175)** | **5987 (12277)** | **0.009** |
| Platelets ($\times 10^9/L$) | 214.5 (95.41) | 214.1 (93.89) | 216.5 (99.51) | 0.703 | **220.6 (94.61)** | **190.6 (95.40)** | **<0.001** |
| Procalcitonin (ng/mL) | 1.41 (7.85) | **0.74 (4.75)** | **2.15 (9.88)** | **0.027** | **0.73 (4.64)** | **3.72 (13.94)** | **0.002** |
| Prothrombin Time (s) | 15.1 (6.2) | 14.8 (7.6) | 15.1 (3.6) | 0.564 | **14.7 (3.8)** | **16.6 (10.9)** | **0.002** |
| RDW (%) | 14.09 (1.98) | 14.08 (2.12) | 13.88 (1.70) | 0.119 | **13.91 (1.96)** | **14.84 (1.93)** | **<0.001** |
| Troponin T (ng/L) | 45.60 (119.6) | **32.38 (92.00)** | **55.04 (138.9)** | **0.009** | **35.17 (106.0)** | **86.66 (156.4)** | **<0.001** |
| WBC ($\times 10^9/L$) | 7.97 (6.96) | **7.22 (6.72)** | **8.87 (7.17)** | **<0.001** | 7.74 (6.37) | 8.85 (8.89) | 0.069 |

Patients coded for “do not intubate” were excluded from analyses for ventilation status. P values are for univariate logistic regression analyses. aPTT = Activated partial thromboplastin time; eGFR = estimated glomerular filtration rate; ESR = Erythrocyte sedimentation rate; MCH = mean corpuscular hemoglobin; MCV = mean corpuscular volume; RDW=red cell distribution width; WBC = white blood cell; SD = standard deviation.

It was notable that age was not a significant predictor of whether a patient would require mechanical ventilation. To investigate this further, we determined rates of mortality and need for mechanical ventilation per decade of life. As anticipated, older age was associated with an increase in mortality rate (**Figure 1a**). However, other than patients in the youngest age groups, the percentage of hospitalized COVID-19 patients requiring mechanical ventilation was similar in each decade of life (**Figure 1b**). Of those that were ventilated, there is a clear correlation between age and risk of death. Indeed, of patients in the oldest group (>84 years of age), only 15% survived if mechanical ventilation was required (**Figure 1c**). Interestingly, young patients were as likely as patients of advanced age to require long durations on ventilation (**Figure 2**). In fact, 78% of ventilated patients between ages 18 and 44 were intubated for longer than 6 days, and 45% were intubated for longer than 14 days.

**Figure 1:**
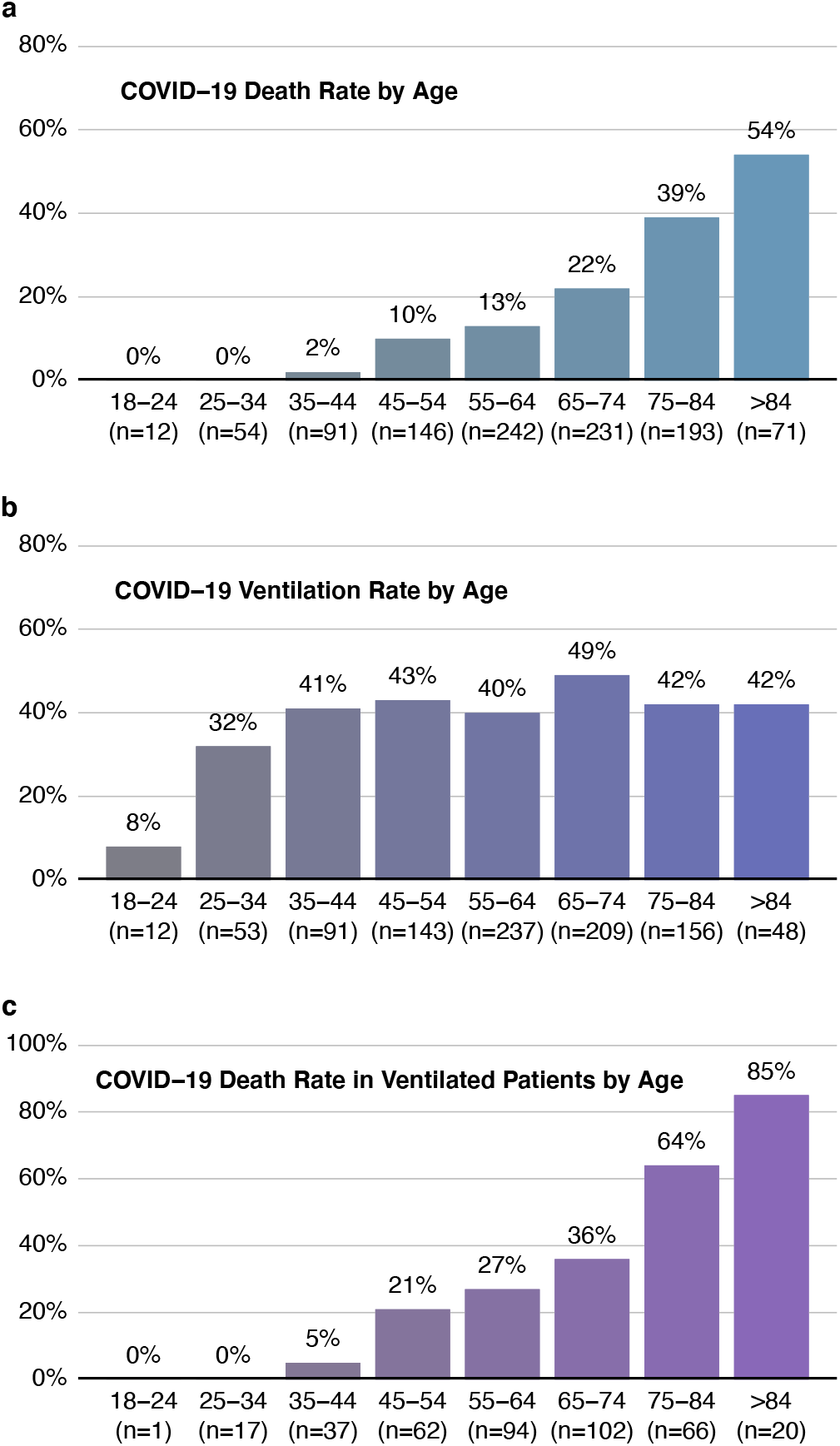
Mortality rate, but not need for mechanical ventilation, increases with age. Mortality rate (a), ventilation rate (b) and mortality rate in ventilated patients (c) were plotted against decade of life.

**Figure 2:**
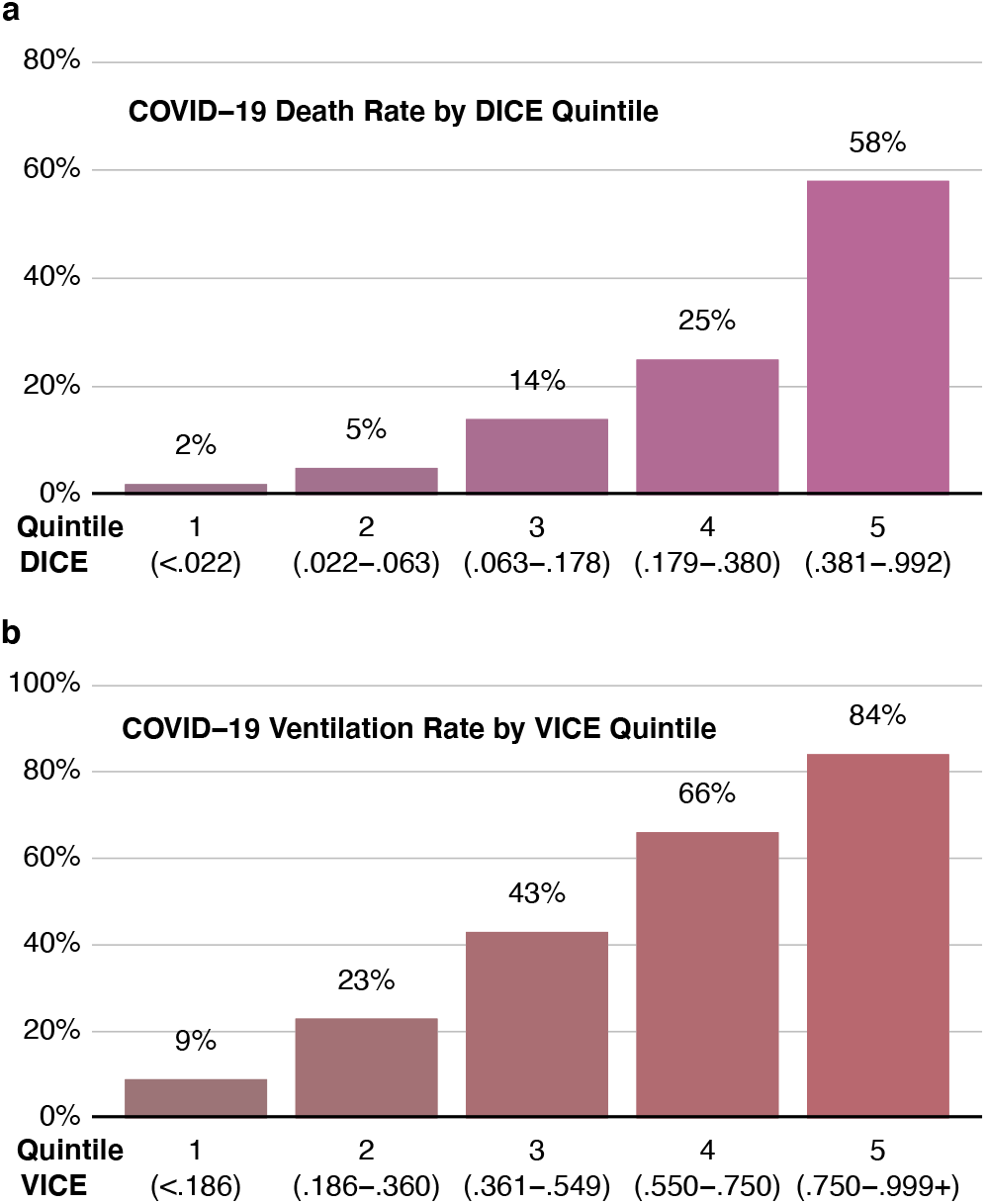
Performance of the VICE and DICE scores in the entire cohort of patients. Ventilation rate was plotted against VICE score quintiles (a). Mortality rate was plotted against DICE quintiles (b).

### Multivariable logistic regression models to predict mechanical ventilation and in-hospital mortality

Multivariable logistic regression models were used in order to develop risk scores to predict important clinical outcomes in COVID-19, namely the need for mechanical ventilation and in-hospital mortality. As detailed in the above section, we found many variables, including age, that were distinctly associated with both the need for ventilation and for mortality. We therefore constructed separate risk scores for ventilation requirement (VICE=Ventilation In COVID Estimate) and death (DICE=Death In COVID Estimate) based on multivariable logistic regression models. We divided our overall study into separate derivation (MGH, n=578) and validation (BWH, NWH, BWFH, NSMC; n=464) populations based on the hospital of admission. Our derivation cohort of 578 patients (**Supplement Table 1**) had a median age of 62 years [IQR, 51-73], consisted of 346 (59.9%) males, and 112 (19.4%) died in the hospital. Excluding patients choosing a DNI status, 242 (45.9%) required mechanical ventilation. Similar to the overall cohort, hypertension (n=304; 52.6%) and diabetes (n=254; 43.9%) were the most common comorbidities.

Using a stepwise backwards regression approach, 7 variables were independently associated with the need for mechanical ventilation (**Table 3**). These variables included diabetes mellitus (OR 1.83; 95% CI 1.17-2.86, P=0.008), dyspnea on presentation (OR 1.76; 95% CI 1.00-3.08, P=0.049), ALT (log2-transformed value, OR 1.30; 95% CI 1.03-1.65, P=0.027), C-reactive protein (log2-transformed value, OR 1.51; 95% CI 1.26-1.81, P<0.001), lactate dehydrogenase (log2-transformed value, OR 2.75; 95% CI 1.75-4.31, P<0.001), neutrophil to lymphocyte ratio (OR 1.45 for every 10-fold increase; 95% CI 1.07-1.99, P=0.018), and troponin-T above 10 ng/mL (OR 1.64; 95% CI 1.02-2.64, P=0.042).

**Table 3:**
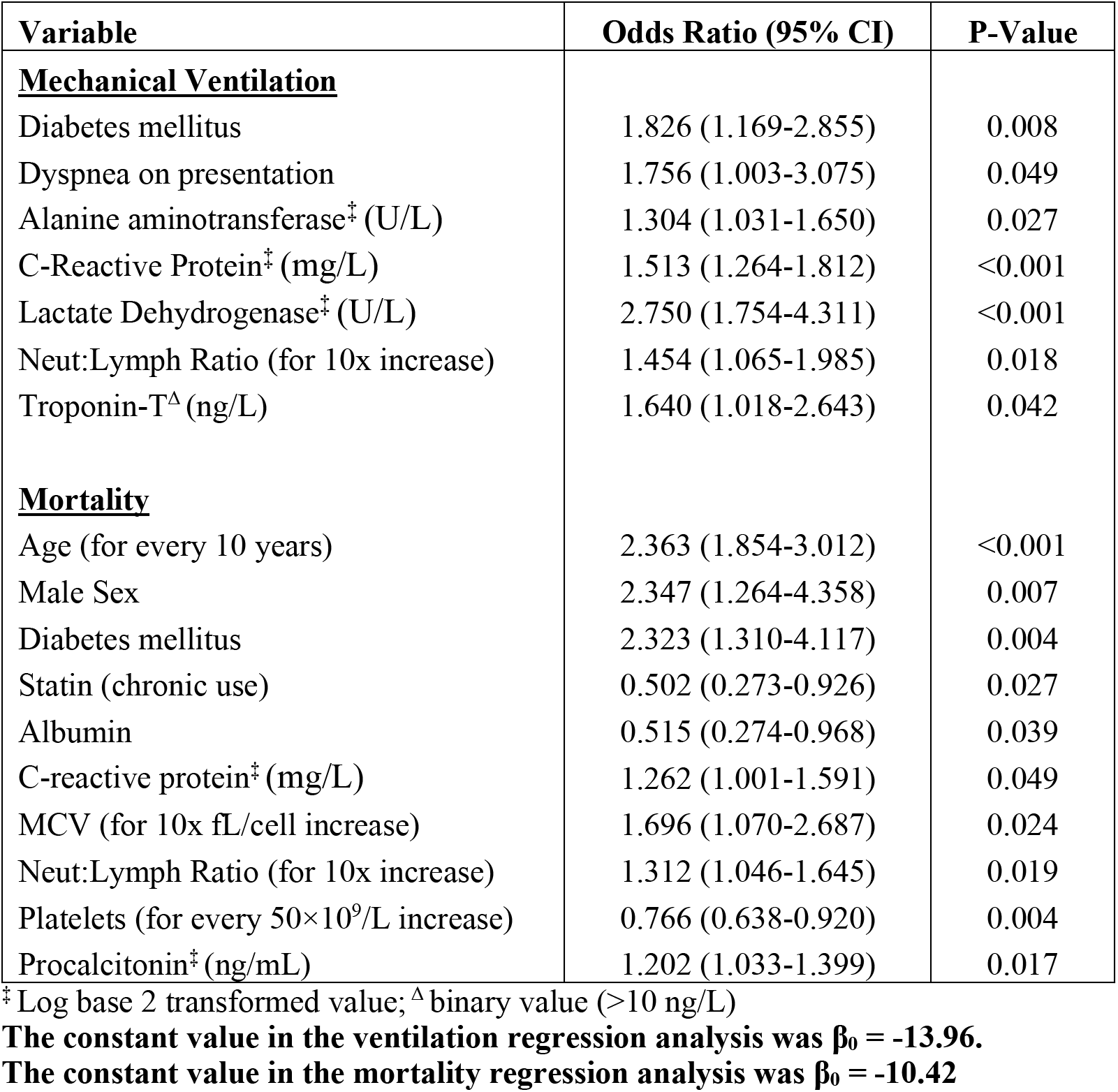
Multivariable logistic regression models for predicting mechanical ventilation need (VICE) and mortality (DICE) in COVID-19 patients.

| Variable | Odds Ratio (95% CI) | P-Value |
| --- | --- | --- |
| **<u>Mechanical Ventilation</u>** |  |  |
| Diabetes mellitus | 1.826 (1.169-2.855) | 0.008 |
| Dyspnea on presentation | 1.756 (1.003-3.075) | 0.049 |
| Alanine aminotransferase<sup>‡</sup> (U/L) | 1.304 (1.031-1.650) | 0.027 |
| C-Reactive Protein<sup>‡</sup> (mg/L) | 1.513 (1.264-1.812) | <0.001 |
| Lactate Dehydrogenase<sup>‡</sup> (U/L) | 2.750 (1.754-4.311) | <0.001 |
| Neut:Lymph Ratio (for 10x increase) | 1.454 (1.065-1.985) | 0.018 |
| Troponin-T<sup>Δ</sup> (ng/L) | 1.640 (1.018-2.643) | 0.042 |
| **<u>Mortality</u>** |  |  |
| Age (for every 10 years) | 2.363 (1.854-3.012) | <0.001 |
| Male Sex | 2.347 (1.264-4.358) | 0.007 |
| Diabetes mellitus | 2.323 (1.310-4.117) | 0.004 |
| Statin (chronic use) | 0.502 (0.273-0.926) | 0.027 |
| Albumin | 0.515 (0.274-0.968) | 0.039 |
| C-reactive protein<sup>‡</sup> (mg/L) | 1.262 (1.001-1.591) | 0.049 |
| MCV (for 10x fL/cell increase) | 1.696 (1.070-2.687) | 0.024 |
| Neut:Lymph Ratio (for 10x increase) | 1.312 (1.046-1.645) | 0.019 |
| Platelets (for every 50×10<sup>9</sup>/L increase) | 0.766 (0.638-0.920) | 0.004 |
| Procalcitonin<sup>‡</sup> (ng/mL) | 1.202 (1.033-1.399) | 0.017 |

<sup>‡</sup> Log base 2 transformed value; <sup>Δ</sup> binary value (>10 ng/L)
The constant value in the ventilation regression analysis was $\beta_0 = -13.96$ .
The constant value in the mortality regression analysis was $\beta_0 = -10.42$

We identified 10 variables (**Table 3**) that independently associated with the odds of death including: age (for every 10 year increase: OR 2.36; 95% CI 1.85-3.01, P<0.001), male sex (OR 2.35; 95% CI 1.26-4.36, P=0.007), diabetes mellitus (OR 2.32; 95% CI 1.31-4.12, P=0.004), chronic statin use (OR 0.50; 95% CI 0.27-0.93, P=0.027), albumin (OR 0.52; 95% CI 0.27-0.97, P=0.039), C-reactive protein (log2-transformed, OR 1.26; 95% CI 1.00-1.59, P=0.049), MCV (OR 1.70; 95% CI 1.07-2.69, P=0.024), neutrophil to lymphocyte ratio (OR 1.31 for every 10-fold increase; 95% CI 1.05-1.65, P=0.019), platelet count (OR 0.77 for every 50 ×10^9^/L increase; 95% CI 0.64-0.92, P=0.004), and procalcitonin (log_2_-transformed, OR 1.20; 95% CI 1.03-1.40, P=0.017). Of note, use of an angiotensin converting enzyme inhibitor (ACEI) or angiotensin receptor blocker (ARB) was not associated with a difference in outcome both in univariate or multivariable analysis.

### Performance of the VICE and DICE risk scores

The VICE and DICE risk scores were constructed based on coefficients from the multivariate logistic regression models. We used the following formula to calculate the probability (*p*) and 95% confidence intervals: 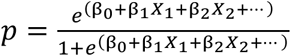 where p = ln(OR) and Po can be found in the caption to **Table 3**. An online calculator based on the VICE and DICE risk scores is available freely to the public, allowing health care providers to enter values for the variables required to calculate the risk for mechanical ventilation and in-hospital mortality at https://covid-calculator.com/. In receiver operator characteristic (ROC) curve analysis, the area under the curve (AUC or C-statistic) of the VICE risk score in the derivation cohort was 0.81 (Supplement **Figure 1a**, 95% CI, 0.77-0.85), and 0.84 (**Supplement Figure 1b**, 95% CI, 0.79-0.88) in the validation cohort. The DICE risk score for mortality in the derivation cohort had an AUC of 0.87 (**Supplement Figure 2a**, 95% CI, 0.83-0.91). The AUC in the validation cohort was 0.80 (95% CI 0.75-0.85) (**Supplement Figure 2b**). As shown in Figure 3, a progressive increase in ventilation (**Figure 3a**) and mortality (**Figure 3b**) rates was observed with increasing quintiles of VICE and DICE scores, respectively, in our cohort. In patients falling within the highest quintile of DICE score, mortality was 58% compared to 2% in the lowest quintile.

**Figure 3:**
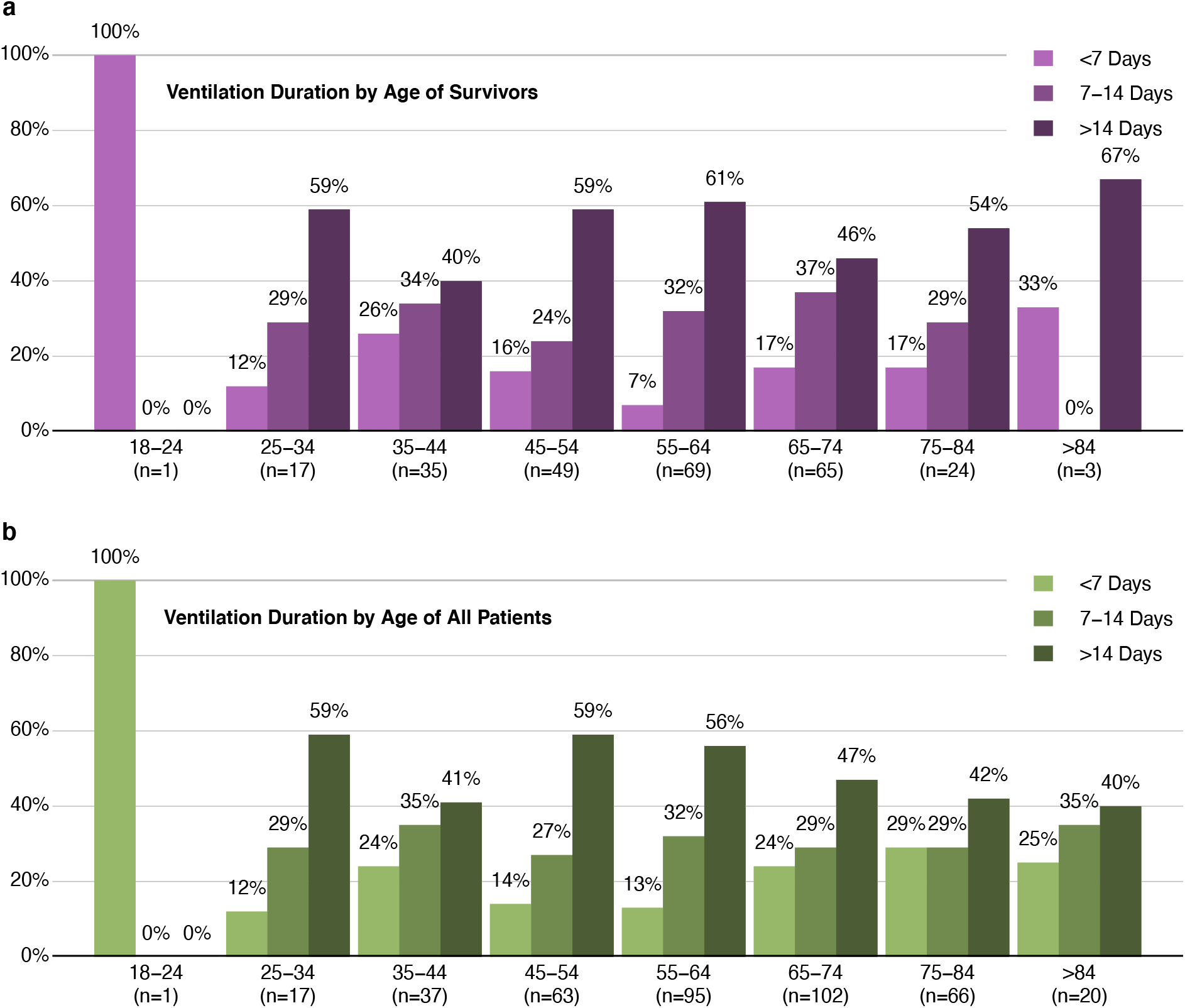
Young people hospitalized with COVID-19 are equally at risk of long ventilation periods as elderly patients. Proportion of length of mechanical ventilation (<7, 7-14, and >14 days) requirement stratified by different age groups in survivors (a) and in the entire cohort (b).

Previous reports have highlighted a marked increase in risk of developing severe illness in COVID-19 in Asians and African Americans.^10-13^ We found it interesting, therefore, that we did not observe this in our univariate analyses (**Table 1**), with the data even pointing to worse mortality rates in white patients. We therefore investigated if patients from white backgrounds were in poorer health on admission to hospital by analyzing their DICE scores. Indeed, we observed that white patients in our cohort were at significantly higher risk of in-hospital death on admission by the DICE score than Black or Hispanic patients (**Supplement Figure 3**, Median DICE Score [IQR] in: White = 0.18 [0.06-0.41)]; Black = 0.12 [0.03-0.30]; Hispanic = 0.06 [0.02-0.19]). Patients from an Asian background also presented with lower risk (Asian = 0.13 [0.03-0.39]), although this did not meet statistical significance. After adjusting for DICE score, race was no longer a predictor of in-hospital mortality. We also considered whether there were a disproportionate number of minority patients in the DNI and/or CMO groups that were excluded from our analyses, therefore explaining the discordant results with the published literature. As shown in Supplemental Table 3, this does not appear to be explained by the CMO population, which had a higher percentage of white patients than any other background. There was also a large percentage of white patients in the DNI group that could have confounded the mortality calculations. However, even when excluding these patients from mortality analyses, Hispanic patients still had a significantly lower risk of death in our study.

Liang et al. recently developed and validated a clinical risk score to predict the development of severe illness (combined end-points of ICU admission, ventilation requirement and death) amongst hospitalized COVID-19 patients.^7^ The algorithm performed very well in cohorts from Hubei (AUC = 0.87) and outside Hubei (AUC = 0.82), but we do not know how well it performs in other populations, including the US. The accuracy of the COVID-GRAM risk score at predicting the combined endpoint of ICU admission, mechanical ventilation requirement, and death in our derivation cohort was 0.71 (0.67-0.76) and 0.70 (0.64-0.75) in our validation cohort. The lower AUC for COVID-GRAM in our population may be due to geographical differences in COVID-19 presentation and outcomes in China compared to Boston.

## Discussion

This study reports on the in-hospital outcomes of sequentially hospitalized patients with COVID-19 in the Boston area. We identified many independent risk factors for mortality in this population, including older age, male sex, preexisting diabetes mellitus, thrombocytopenia, hypoalbuminemia, and higher levels of inflammatory and infectious biomarkers including procalcitonin, CRP, and neutrophil to lymphocyte ratio. Interestingly, we also found that chronic statin use was associated with a lower risk of death, perhaps supporting the anti-inflammatory and immunomodulatory benefits of statins in this disease.^14-16^ Notably, there was some but not complete overlap in the factors that predicted ventilation needs, with preexisting diabetes and elevated CRP and neutrophil to lymphocyte ratio being included in both risk score models.

Factors that uniquely and independently predicted ventilation requirement included dyspnea on presentation and elevated lactate dehydrogenase, ALT, and troponin. Age was not a significant predictor of ventilation need, perhaps dispelling the belief that COVID-19 only severely affects the elderly. Recent evidence suggests that young people are driving the recent surge in coronavirus cases in many states. While there is clear evidence that young patients are less likely to die from COVID-19, young hospitalized patients regularly require the use of ventilators for extended periods of time, thus stressing a system that is already in short supply of ICU beds. Recognizing the difference in variables that independently predict need for ventilation and death in COVID-19 is greatly important, especially as current risk score calculators combine ICU admission, ventilation needs and mortality into one endpoint.^7^ The use of two independent risk scores predicting ventilation need and death allows health care systems to not only more precisely prognosticate individual patient outcomes but also better predict demand for mechanical ventilators and ICU beds.

In our study, we demonstrated that excessive levels of inflammatory and infectious markers, such as CRP and procalcitonin, were associated with an increased risk of death. Given the relationship between cardiometabolic disease and death in COVID-19, it is possible that these patients are more vulnerable to the aggressive inflammatory response induced by the virus.^17^ Of note, it has recently been suggested that biological, rather than chronological, aging is a stronger predictor of all-cause mortality related to COVID-19.^18^ Therefore, underlying age-related cardiovascular dysfunction may increase the risk of a hyperinflammatory response that augments the effects of COVID-19 in these individuals.^17,19,20^ Interestingly, elevated troponin levels, a marker of cardiac injury, predicted increased need for mechanical ventilation in multivariable analysis whereas chronic statin use was associated with reduced in-hospital mortality, further underscoring the strong link between underlying cardiovascular disease and worse outcomes with COVID-19.^8,21-29^

Importantly, we developed two novel prediction models to calculate risk of hospitalized COVID-19 developing severe outcomes. These models were effective at predicting risk of ventilation need and death in both our derivation and validation cohorts. Of note, the two cohorts showed considerable variability in several factors, including sex and age, demonstrating our risk scores may be accurate in distinct populations from the United States. We also observed that the factors used to construct each risk score were different, demonstrating that it is beneficial to consider risks of ventilation need and mortality separately. Interestingly, and in contrast to previous reports, we did not observe worse outcomes in COVID-19 patients from minority ethnic backgrounds ^10-13^ In fact, we actually presented data suggesting worse mortality rates in white patients, even when removing patients with a DNI status. However, we found no difference in outcomes after adjusting for individual DICE scores. These findings suggest that variations in outcomes by racial group may be explained by differences in underlying risk factor profiles, although socioeconomic status was not assessed in our study and could be another determinant of outcome. In addition, the patients included were inpatients only, and inequitable access to healthcare (including hospital admission) appears to contribute to the disproportionate impact of COVID-19 on patients from racial minorities in the United States.^30^ The data from our study was unable to quantify any racial disparities in this other important area.

The variables that we found to predict mechanical ventilation requirement and mortality are either readily available or routinely measured upon admission to the hospital. We anticipate that clinicians will easily be able to implement the DICE and VICE scores to stratify risk in admitted patients. In situations where hospital resources are plentiful, clinicians could use both scores to identify which patients are most likely to develop severe illness, and plan accordingly to monitor higher risk patients more intensely. However, under the most critical of circumstances where ventilators are in short supply, clinicians may require aid in triage and ventilator utilization. For example, ventilators may be prioritized for those patients who are at most risk for ventilation need (high VICE score) while still having a relatively lower risk of death (assessed by the DICE score).

One of the main strengths of the study was our ability to collect comprehensive data on and follow more than 99% of the patients from our cohort from admission to the primary endpoint, either discharge or death, with a large fraction of patients (18%) remaining in the hospital for longer than 28 days. Also, data were obtained by detailed medical record review rather than reliance on billing codes. Given the different variables that predict ventilation needs and mortality, we believe another strength was the construction of distinct risk scores. One potential limitation is the modest sample sizes in both our derivation and validation cohorts. Although the exclusion of CMO patients was justified in the current study, this may have resulted in underestimating the impact of malignancy on inpatient mortality. Further, we aimed to focus on admission findings in determining outcomes and hence our risk scores do not include the effects of different treatment regimens.

This study identified baseline patient characteristics and admission laboratory values that associate with critical illness and in-hospital death in patients with COVID-19. In this investigation, we developed and validated risk score calculators to predict mechanical ventilation need and in-hospital death in COVID-19 patients. These risk scores could potentially aid clinicians to better stratify risk in COVID-19 patients and optimize patient care and resource utilization in the surge of infections we are facing worldwide.

## Data Availability

All data are presented in the paper in summary statistics. Additional data will be made available upon request of the corresponding author.

## Authorship contributions

CJN and RM conceived, designed, and planned the study with input from LW and HHS. WT, WJ, RHL, HHS, and LW extracted the data from RPDR and EPIC. LW and HHS designed the code that was used to extract data from RPDR and analyze the data. CJN, HHS and LW performed the data analysis and designed the figures. CJN, LW and RM interpreted the data. CJN and RM drafted the manuscript with critical input from CLLC, LW, HHS and RHL. All authors interpreted the data and made significant contributions to manuscript editing.

## Acknowledgements

Drs. Nicholson and Malhotra are supported by a COVID-19 Fast Grant (fastgrants.org). Dr. Malhotra is supported by NHLBI R01 HL142809, the American Heart Association grant 18TPA34230025, and the Wild Family Foundation. Dr. Lino Cardenas is supported by the MGH Physician-Scientist Development Award and the MGH Department of Medicine Pilot Translational Research Grant.

